# Probing the Surgical Competence of LLMs: A global health study leveraging AfriMedQA benchmarks

**DOI:** 10.1101/2025.10.05.25337350

**Authors:** Tobi Olatunji, Folafunmi Omofoye, Ezinwanne C. Aka, Gina Itzikowitz, Daniel Macaulay, OyinOluwa G. Adaramola, Boluwatife A. Adewale, Chidi Asuzu, Simisola Popoola, Wendy Kinara, Emmanuel Ayodele, Ifeoluwa Yinusa, Oluwatoni Adekunle, Mardhiyah Sanni, Chibuzor Okocha, Tassallah Abdullahi, Abraham Owoduni, Charles Nimo, Mercy N. Asiedu, Bilal Mateen, Rebecca Weintraub

## Abstract

Global surgical care faces a severe workforce shortage, with more than 1.2 million additional specialists needed by 2030, particularly in low- and middle-income countries (LMICs). Large language models (LLMs) have demonstrated impressive medical reasoning on standardized exams, but their safety, reliability, and specialty-specific performance—especially in procedural fields such as surgery—remain uncertain. Here we evaluate over 40 state-of-the-art LLMs on 3,900 expert-authored multiple-choice questions across 32 medical specialties from the AfriMed-QA benchmark, developed by 20 African medical professors. Top models (o1, GPT-4o, Claude 3.5) achieved mean accuracies exceeding 82%, showing strong diagnostic reasoning, yet consistently underperformed in surgery, pathology, and obstetrics compared with medical disciplines. Error analyses revealed frequent procedural reasoning failures, omission of local clinical guidelines, and overconfident but incorrect answers. Smaller or biomedical models exhibited higher hallucination and formatting error rates, while prompting strategies had inconsistent benefits. These results highlight the uneven readiness of LLMs for specialty-specific decision support and underscore the need for locally grounded evaluation frameworks, improved instruction tuning, and rigorous real-world validation to ensure the safe and equitable deployment of AI-assisted clinical tools in LMICs.

## Introduction

The global surgical workforce is facing an unprecedented shortfall. By 2030, an estimated 1.27 million additional surgeons, anesthetists, and obstetricians will be required to achieve minimum surgical service coverage worldwide, with the most severe shortages concentrated in low- and middle-income countries (LMICs) ^1–3^. In Sub-Saharan Africa, wait times for essential and elective surgical care continue to rise as the availability of specialists lags behind population growth and the increasing burden of disease ^4,5^. This persistent shortage has led to urgent calls for scalable, technology-enabled solutions to help bridge workforce gaps and support clinical decision-making across diverse care settings ^5,6^.

Recent advances in large language models (LLMs) present a promising opportunity. Models such as GPT-4, Claude-3.5, and Med-PaLM have demonstrated exceptional capabilities in medical reasoning tasks, including clinical summarization, diagnostic support, and question answering ^7–11^. In controlled evaluations, models like GPT-4 have outperformed human clinicians on USMLE-style medical licensing examinations ^10–14^. This “superhuman” performance has generated growing interest in leveraging LLMs to augment care delivery, particularly in resource-constrained settings ^9,15^.

However, the credibility of such evaluations has come under scrutiny due to benchmark contamination, wherein questions from publicly available datasets are inadvertently included in model pre-training corpora ^16^. To address this issue, newer benchmark initiatives such as AfriMed-QA, CMExam, and MedQA-SWE have emerged, offering regionally diverse, specialty-rich, and previously unseen datasets to test LLM generalizability in real-world clinical scenarios ^16–18^. This focus on generalizability is especially critical for LMICs, where clinical contexts, disease burdens, and guideline frameworks often differ significantly from those in high-income countries. Ensuring model performance in these settings is essential not only for equity but also for the safety, effectiveness, and trustworthiness of AI-assisted clinical decision support.

AfriMed-QA, for example, comprises over 15,000 questions sourced from more than 60 medical schools across 16 African countries and 32 clinical specialties, including surgery, pediatrics, and obstetrics & gynecology ^16^. This dataset enables a robust evaluation of model performance across linguistic, geographic, and clinical contexts that remain underrepresented in most global LLM training corpora ^16^. In these more rigorous and diverse settings, top-performing general-purpose models like GPT-4o and Claude 3.5 have continued to perform well, while smaller biomedical-specific models have lagged, often exhibiting hallucinations and issuing potentially harmful recommendations in high-stake specialties like surgery and infectious diseases ^8,16^. These trends carry significant implications for LLM deployment in LMICs, where surgical capacity is particularly limited and the need for trustworthy decision support tools is especially urgent ^19,20^.

Despite these challenges, LLMs continue to outperform human clinicians in consumer-facing tasks, such as patient education and basic triage advice, with blinded raters preferring LLM responses for their completeness and clarity ^21^,^22^. However, using LLMs for specialty decision support—especially in high-risk areas such as surgery—requires a deeper look into how and when errors occur, including their frequency, nature, and potential clinical impact.

This study explores how large language models (LLMs) perform in surgical specialties using vignette-style clinical multiple-choice questions (MCQs) from the AfriMed-QA benchmark. Our core research question is: **How do the types and frequency of LLM errors differ between surgical and non-surgical specialties**? By examining accuracy, omissions, hallucinations, and potential for harm, we aim to support the safe, fair, and effective use of LLMs in surgical care worldwide.

## Results

**Figure 1:**
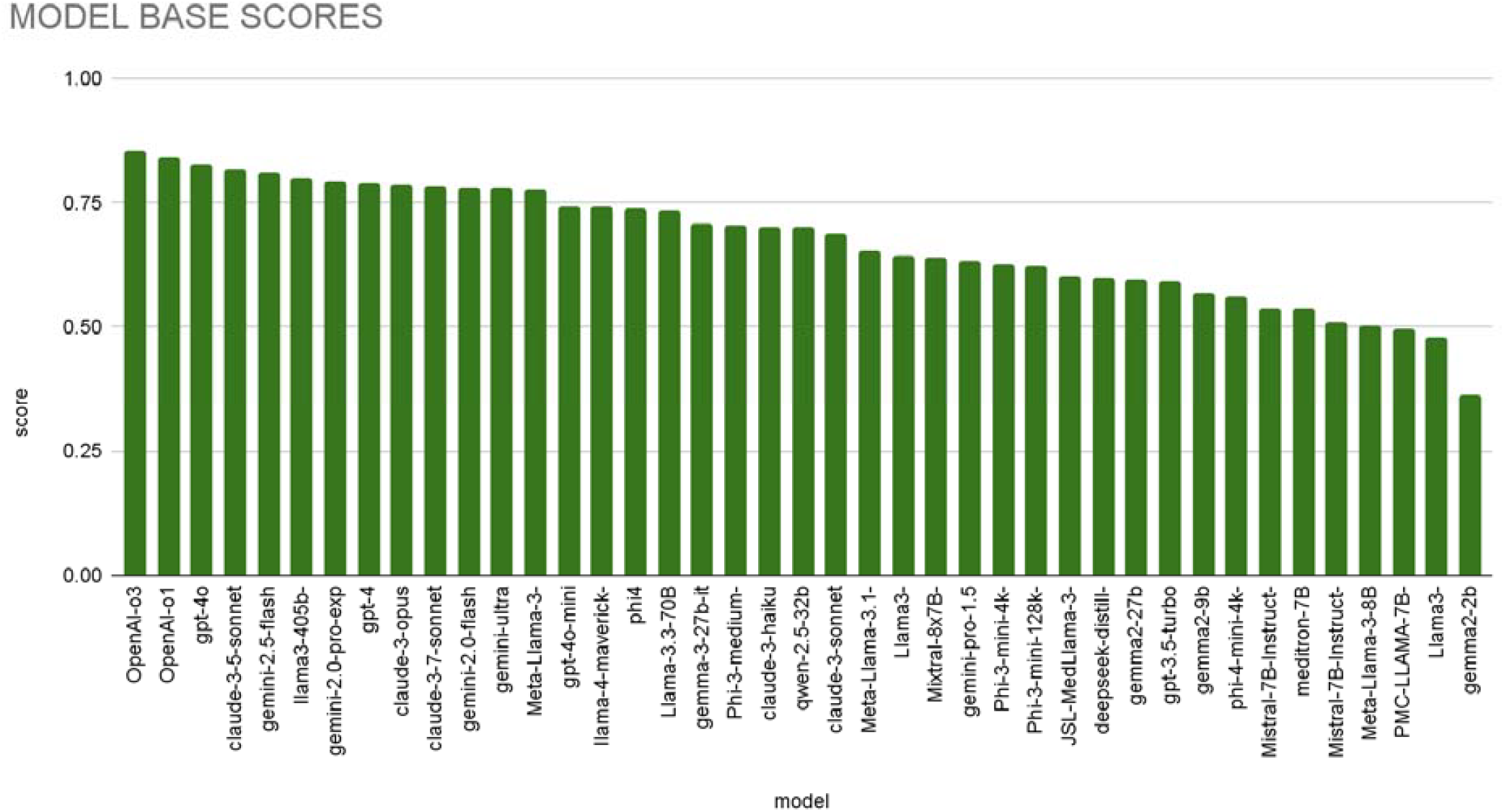
Expert MCQ accuracy across 42 LLMs

### Overall LLM Performance on Expert MCQs

We evaluated 42 LLMs (9 recently released LLMs) on 3,910 expert-written multiple-choice questions (MCQs) from African countries spanning 30 medical specialties. Overall, newer models outperformed earlier variants. The top-performing model was **OpenAI-o1**, achieving an accuracy of **84.2%**, replacing **GPT-4o (82.9%)**, the previous best model. Gemini-2.0 models (pro and flash) surpassed earlier generations (Ultra/1.5-pro). Gemma-3 surpassed Gemma-2, Phi4 surpassed Phi-3 variants. **LLaMA3-405 B-Instruct-MAAS (80.1%)** retained the best open model position.

**Figure 2:**
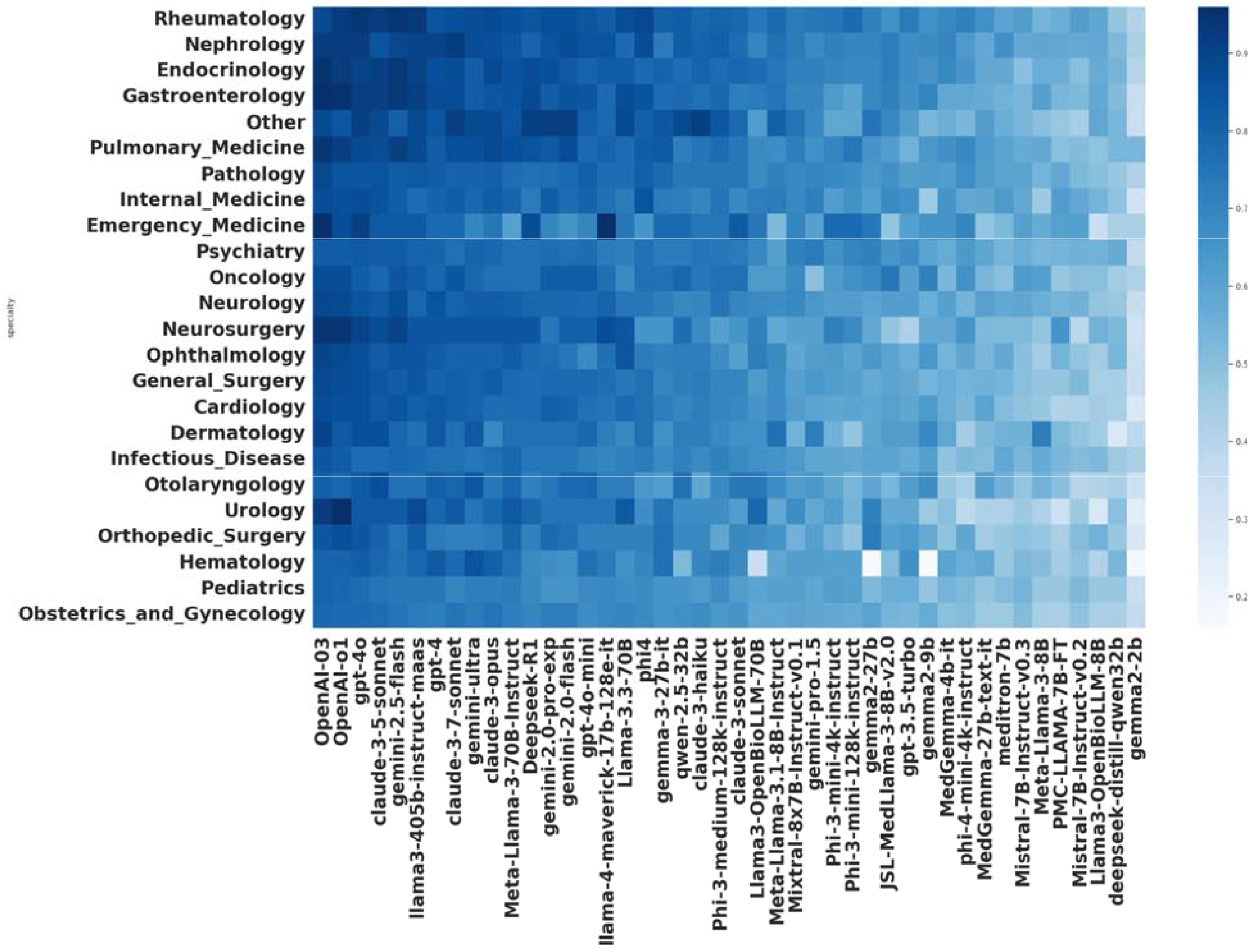
Accuracy scores for 43 LLMs on Expert MCQs across 24 specialties

### Performance Across Medical Specialties

When disaggregated by specialty, model accuracy varied widely. Most models performed well in **internal medicine, endocrinology**, and **rheumatology**, achieving accuracies above 85%. However, accuracy dropped sharply in **surgery, pediatrics, obstetrics & gynecology**, and **pathology**—specialties requiring procedural reasoning or context-aware clinical judgment. For instance, Claude-3.5-Sonnet achieved 92% in rheumatology but only 81% in surgery and 78% in pediatric surgery.

**Figure 3:**
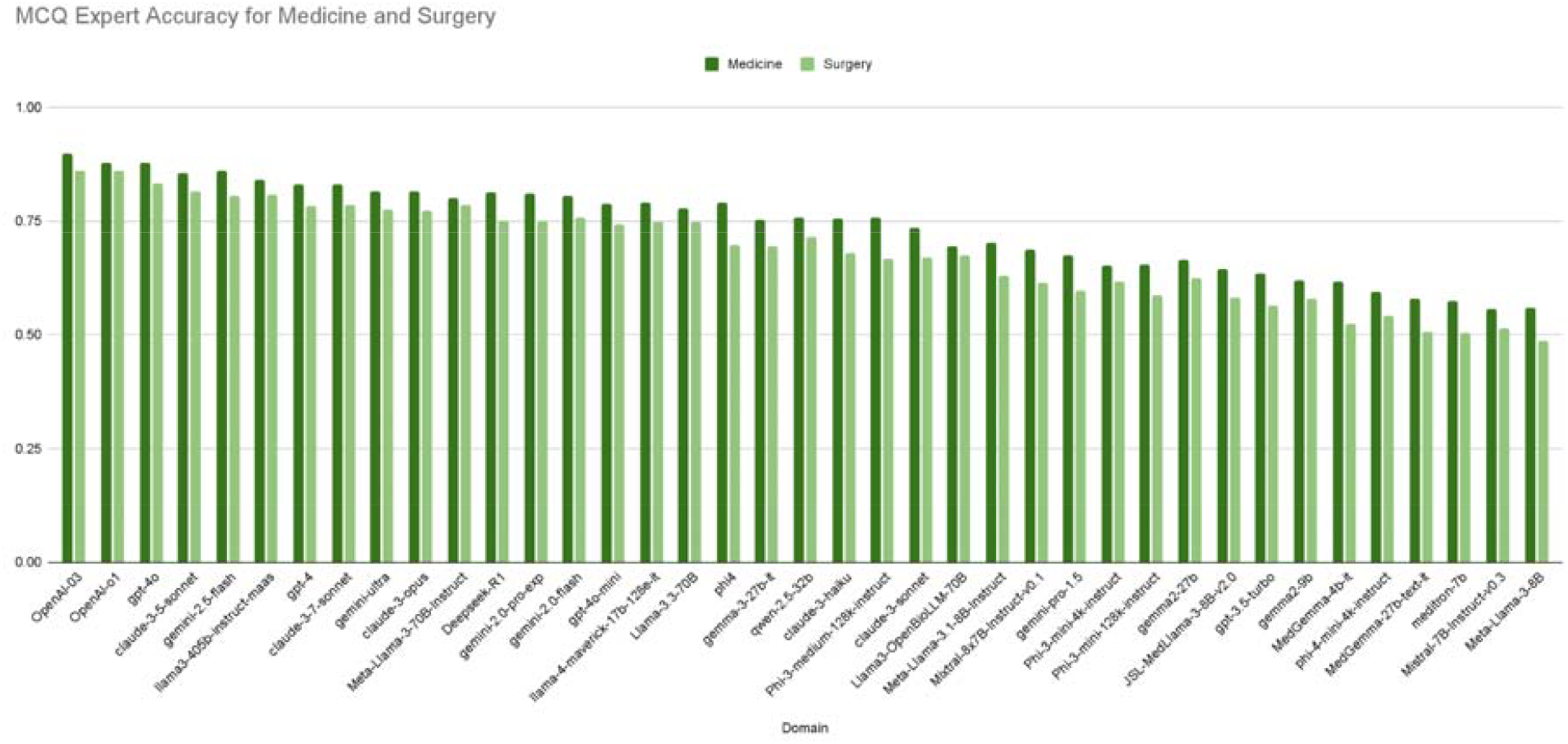
Accuracy scores for 42 LLMs on Expert MCQs from Medical, Surgical specialties

### Surgical vs Non-Surgical Specialties

We aggregated specialty performance into two major categories–surgical (e.g., general surgery, orthopedic surgery, OB/GYN) and Medical (e.g., cardiology, infectious diseases). Across all models, **Medical specialties outperformed surgical specialties by 5–10 percentage points** on average. For GPT-4o, the accuracy was 87.7% in medicine vs. 83.2% in surgery; Claude-3.5-Sonnet similarly scored 85.6% in medicine and 81.4% in surgery.

A statistical comparison (T-test) confirmed the difference between Medical and Surgical specialty performance to be highly significant (p < 0.0000001 across models), suggesting that surgical and procedural domains present distinct challenges to LLMs.

**Figure 4:**
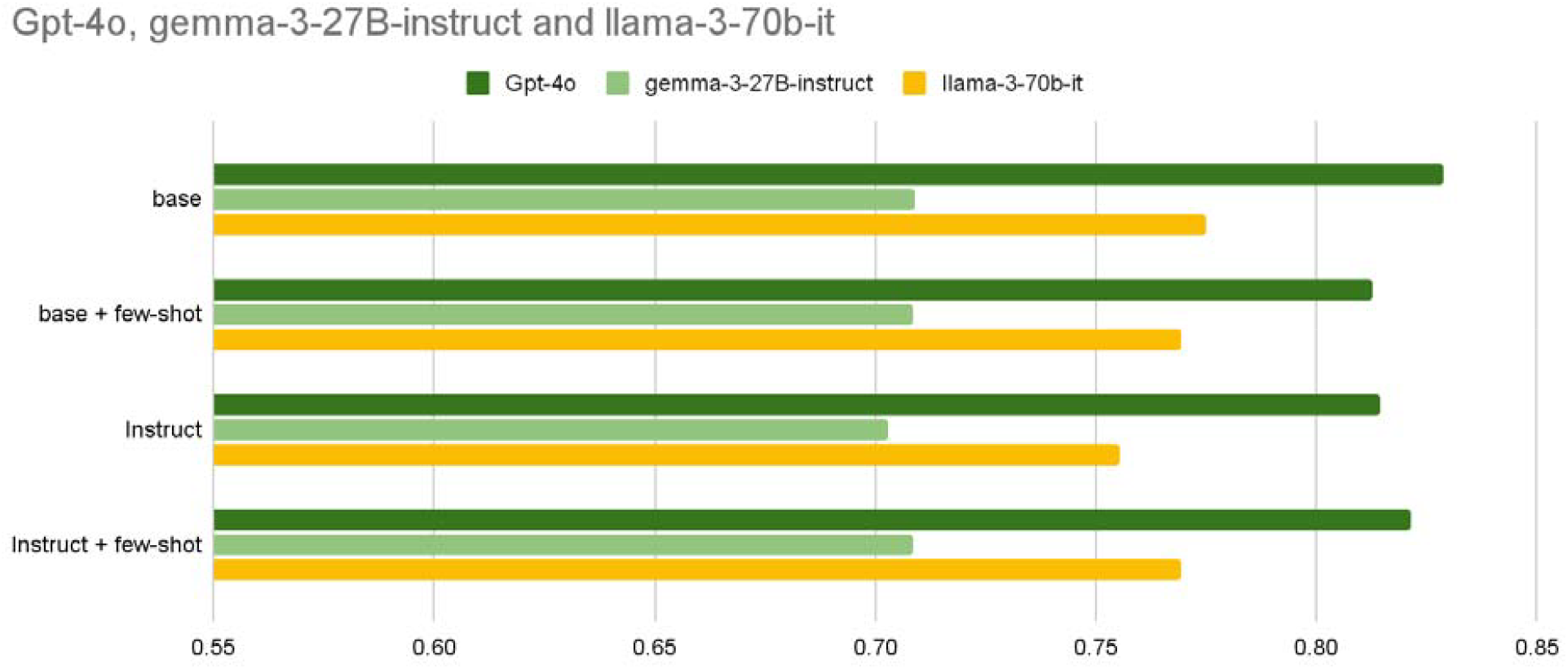
Effect of prompting strategies on MCQ accuracy of selected LLMs

### Prompting Strategies: Base vs Instruct vs Few-Shot

Prompt engineering had an unexpected mixed and muted effect on model accuracy, with base prompts outperforming “instruct” and “few-shot” prompting for open and closed models, as well as large, medium, and small models evaluated. This requires further investigation. Interestingly, instruct+few-shot consistently outperformed instruct prompting, indicating prompting techniques could yield some benefit.

**Figure 5:**
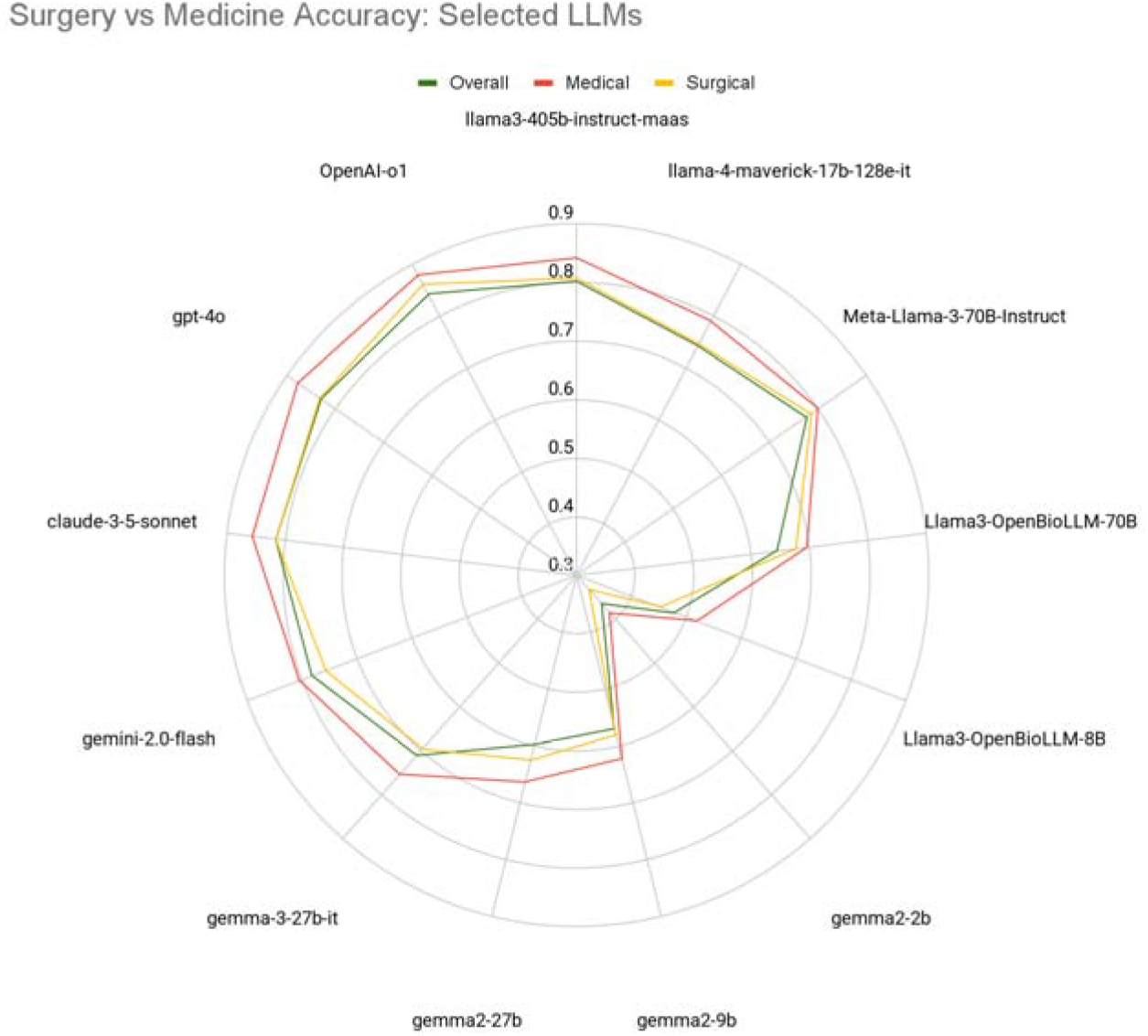
Expert MCQ performance across selected large, small, open, closed, general, and biomedical LLMs

### Closed vs Open Models

Closed-source models (e.g., OpenAI, Anthropic, Google) outperformed open models by a wide margin, and this trend was consistent across Medical and Surgical specialties. The top three models were all proprietary, with an average score of 87% for Medicine and 83.5% for Surgery, compared to 81% and 78% for the top 3 open models. Among open models, the most competitive were **Llama3-405B, Meta-Llama-3-70B-Instruct**, and **Llama-4-maverick**, all by Meta.

### Model Size and Architecture

Model size correlated strongly with performance. Models with >30B parameters consistently scored 10–15 percentage points higher than smaller counterparts, and this trend was consistent across Medical and Surgical specialties. This pattern is evident with model families or variants of different sizes, for example, Gemma-2 variants, Llama-3 variants, GPT variants, and Gemini variants, where larger model variants consistently outperform smaller (or distilled) variants trained on similar datasets with similar architecture.

### Effect of Training Data and Training/Tuning Approach

Gemma-2-27B (Med=0.66, Surg=0.62) and Gemma-3-27B (Med=0.75, 0.69), one generation apart, with the same number of parameters, with nearly 10 points difference in performance, demonstrate that model size/architecture are not the only performance drivers. Although the accuracy improvement in Surgery (0.07) still lags Medical specialties (0.09), improved data and training approaches for later generation models yield performance gains at constant parameter size.

### General vs Biomedical Domain Finetuning

Surprisingly, across Medical and Surgical specialties, **biomedical-specialized models** such as **Meditron-7B, OpenBioLLM-8B**, and **PMC-LLAMA-7B** underperformed compared to general-purpose LLMs. While these models occasionally performed well on isolated pathology questions, they struggled with reasoning, context interpretation, and question formatting, suggesting that biomedical pretraining alone is insufficient.

### Manual Error Analysis

Manual review of correct vs incorrect responses across six representative models revealed that errors in **surgical specialties** were more likely to involve:

**Table 2:** Results of manual error coding showing counts of each error type.

|  |  | 125 Total Errors<br>manually reviewed by experts (Pilot 1 Results) |  |
| --- | --- | --- | --- |
| Error Code | Description | 79 Surgical Domain Errors<br>(Obs&Gyn + Gen Surg + Peds Surg) | 46 Medical Domain Errors<br>(Internal Medicine + Infectious Dx)) |
| E1 | Factually Inaccuracies | 37 | 16 |
| E2 | Hallucinations | 4 | 4 |
| E3 | Outdated Information | 1 | 0 |
| E4 | Failure to follow local Medical / Population-Specific guidelines | 6 | 4 |
| E5 | Invalid Inference | 17 | 4 |
| E6 | Incorrect Surgical-Clinical Reasoning | 0 | 0 |
| E7 | Inappropriate Management | 4 | 0 |
| E0 | Prompting | 10 | 18 |

#### Manual Error Code Analysis Count by Expert Panel

Manual error analysis revealed a range of examples of questions the models struggled with, including: surgical vignettes in which next best management questions were asked, to pediatric management questions where the age range (6 months to 1 year vs 1 to 6 years etc) would impact the clinical decision making. Models also struggled with surgical questions that might require local context, ex, “commonest cause of cellulitis in Africans” being different to the generally agreed answer outside of Africa, highlighting the importance of incorporating regional epidemiology into LLM training. Further, LLMs frequently defaulted to guidelines or disease patterns typical of high-income countries, missing nuances relevant in African settings. Interestingly, models often performed well on pharmacological reasoning tasks that required understanding the mechanism of action or receptor-level effects of drugs.

Initial counterfactual testing across 4 high-performing LLMs (GPT-4o, Gemini, Meta AI, DeepSeek) reveal a <1% occurrence of an inappropriate change in LLM answer when patient variables like age, gender, or geographic location were altered in the clinical question for the purpose of assessing implicit bias, and susceptibility to demographic shifts. Results from initial counterfactual testing are encouraging but will require further, scalable evaluations.

## Discussion

### Statement of Principal Findings

This study presents a comprehensive evaluation of 42 large language models (LLMs) on a diverse set of clinical questions sourced from African medical educators, with a focus on performance variation across surgical and non-surgical specialties. Our findings demonstrate that state-of-the-art models such as o1, GPT-4o, Claude-3-5, and Gemini Ultra achieve >80% accuracy, exhibiting impressive reasoning capabilities across a wide range of clinical specialties, a tough task for most clinicians, reinforcing their potential utility in decision-support roles.

However, significant performance disparities persist between specialties. Surgical specialties, along with Obstetrics & Gynecology and Pathology, consistently yielded lower LLM accuracy compared to specialties like Internal Medicine, Rheumatology, and Endocrinology. This variance has critical implications for LLM deployment in global health, particularly in domains where decision-making complexity and patient safety risks are highest.

### Mechanisms and Explanations

LLMs consistently underperformed in surgical specialties, where accurate reasoning often requires integrating anatomy, procedural sequences, and clinical management pathways—skills that may not be adequately captured during pretraining on general web and biomedical corpora. Many errors in these specialties involved plausible but incorrect reasoning, often on questions specifically designed to test careful interpretation or sequential logic. For instance, distractor-heavy surgery questions confused even top models, particularly when they required distinguishing between closely related steps in patient management.

In pediatric surgery, most model responses were outright incorrect. This may reflect not only limited exposure to pediatric-specific training data but also a failure to adjust reasoning based on age-dependent variations in disease presentation and treatment guidelines.

Models frequently produced overconfident but incorrect answers, generating detailed rationales that appeared persuasive but contradicted clinical best practices. These were especially concerning in high-stakes contexts, such as decisions about antibiotic prophylaxis or surgical interventions in infants.

Further, models showed limited awareness of local guidelines, often defaulting to global standards inappropriate for African clinical settings. For example, models failed to recommend penicillin prophylaxis for Streptococcus pyogenes in rheumatic heart disease, or to incorporate transfusion protocols for sickle cell crises, suggesting a gap in context-sensitive reasoning essential for global health deployment.

Finally, models struggled with second-order reasoning, particularly when questions required understanding the downstream effects of clinical decisions in resource-limited environments (e.g., choosing a cesarean section in a setting with poor access to surgical follow-up).

### Strengths and Weaknesses of the Study

A key strength of our study lies in the use of the **AfriMed-QA expert subset**—a geographically diverse, specialty-rich, and previously unseen benchmark that avoids contamination from training corpora. Unlike prior work that relied heavily on USMLE-style test banks ^11,13,23^, our dataset reflects real-world LMIC clinical scenarios, authored by practicing African physicians. This enhances external validity and provides more meaningful insight into global model generalizability.

The inclusion of a wide array of model architectures (open vs. closed, general vs. biomedical, large vs. small) allowed us to explore performance differences under realistic constraints. However, limitations include the focus on multiple-choice and short-answer formats, which may not fully capture the nuances of real-time clinical decision-making. Additionally, while human evaluation was incorporated, it was not scaled across all 40+ models due to resource constraints.

Our evaluation focused on structured Multiple Choice Questions (MCQs) and Short Answer Questions (SAQs), which do not capture the full complexity of clinical encounters. Biomedical models were underrepresented among the top performers, pointing to gaps in current domain adaptation strategies. Finally, while we examined error patterns and prompting effects, further investigation is needed to assess real-world impact, trust calibration, and long-term model behavior in dynamic care settings.

### Comparison to Prior Work

Our findings build upon previous studies that demonstrated LLM success on Western-centric medical benchmarks like MedQA and MultiMedQA ^23^. However, unlike these prior evaluations, we highlight a more granular specialty-level breakdown, exposing key performance gaps in high-risk domains. For instance, GPT-4o and Claude-3.5, while excelling in Internal Medicine, struggled with Pediatric Surgery, Anatomy, and Contextual Management in Surgery—mirroring trends reported in localized evaluations such as MedQA-SWE ^24,25^.

### Common LLM Issues Observed

Several systemic issues were observed across LLMs:

- **Hallucinations** were common in smaller, open models (e.g., Meditron-7B, Phi-3 Mini, BioMistral-7B), especially when faced with ambiguous or multi-layered questions.
- **Formatting inconsistencies** and verbosity were also more frequent in these models, complicating their use in structured EHR or exam settings. **Overanalysis** occurred in several models, leading to the rejection of correct answers in favor of elaborate yet incorrect logic.
- A subtle **bias toward certain MCQ options** (e.g., favoring choice “C”) was noted across multiple runs, echoing patterns observed in recent evaluations[Zhang et al., 2023].
- Notably, **LLM scale** was the most consistent predictor of performance. Larger models (>30B) significantly outperformed smaller models, though well-prompted small models could narrow the gap.
- Finally, **domain-specific finetuning alone was insufficient**: biomedical models like PMC-LLAMA-7B and Meditron-7B underperformed across most specialties, highlighting the importance of both scale and instruction tuning.

The accuracy drop seen in surgery, pediatrics, obstetrics & gynecology, and pathology continues to point to weaknesses of current models at reasoning tasks that involve nuanced clinical reasoning as opposed to general questions requiring pure recall.

Errors in non-surgical specialties were more often due to subtle omissions or ambiguities in differential diagnosis. Overall, surgical questions demanded higher precision in context recognition, procedure selection, and risk assessment.

Similar findings are documented when analyzing LLMs’ mistakes in a clinical context, with common issues found to include factual inaccuracies, especially when questions were complex or detailed ^26,27^.

These insights underscore the need for specialty-specific testing and post-deployment error surveillance when using LLMs for clinical support.

Model performance was also influenced by architecture and training scale. Closed models with extensive reinforcement learning and instruction tuning outperformed smaller open models by large margins. Contrary to expectation, biomedical-specific models such as Meditron-7B and PMC-LLAMA-7B did not consistently outperform generalist models, likely due to narrower domain pretraining and reduced parameter scale.

### Implications for Clinicians and Policymakers

The rapid advancement of LLMs in clinical reasoning opens new opportunities for scaling surgical expertise and supporting frontline clinicians, particularly in LMICs. While high-performing LLMs could offer scalable support for clinical education, documentation, and even frontline care, their uneven performance across specialties and geographies must be addressed before widespread adoption.

Our findings highlight the importance of specialty-specific validation and local contextualization before clinical deployment. Models must be benchmarked not only on generic performance but also on error typology and real-world consequences in high-risk specialties.

For clinicians, LLMs may become valuable assistants in history-taking, triage, and documentation—but not yet in autonomous decision-making, especially in procedural care or pediatric settings.

For policymakers, the findings underscore the need for regulatory pathways and clinical trials that emphasize performance by specialty and geography. Investing in local dataset creation, like AfriMed-QA, and incorporating regional guidelines into LLM fine-tuning pipelines will be essential to ensuring safe and equitable deployment. Post-deployment surveillance frameworks are also critical to detect and mitigate hallucinations or unsafe suggestions.

### Unanswered Questions and Future Research

Several questions remain. How can LLMs be trained or fine-tuned to better represent procedural knowledge? What is the optimal prompting strategy for real-time surgical decision support? Can LLMs be made to reason effectively across heterogeneous health systems with minimal bias? Further work is needed to develop post-deployment monitoring frameworks, explore multimodal inputs (e.g., combining imaging with clinical text), and evaluate LLMs in simulated care settings with human-in-the-loop decision-making. Further interdisciplinary work is urgently needed to move from evaluation to safe, equitable, and impactful deployment of LLMs in global health.

## Conclusion

This study presents the most extensive specialty-level evaluation to date of large language models (LLMs) on clinically-authored questions from low- and middle-income countries (LMICs), with a particular focus on surgical domains. Leveraging the expert subset of the AfriMed-QA benchmark, we assessed over 40 LLMs across surgical and non-surgical specialties, uncovering key patterns in model accuracy, error types, and the influence of prompting, model scale, and domain specialization.

Our findings confirm that state-of-the-art models such as GPT-4o and Claude-3-5 achieve superhuman performance in several specialties, demonstrating significant potential for augmenting clinical decision-making and improving access to specialty knowledge in regions facing acute healthcare workforce shortages. At the same time, we identify important limitations in current LLM capabilities—particularly in surgical, pediatric, and context-sensitive domains— where models struggle with procedural reasoning, factual recall, and local guideline adherence.

The implications for society are profound. If carefully validated and deployed, LLMs could offer scalable decision support tools to address the global shortfall in surgical expertise, accelerate medical training, and expand equitable access to high-quality care. For the research community, our work highlights the importance of geographic and specialty diversity in LLM benchmarks and the need for evaluations that go beyond aggregate scores to scrutinize error types, safety, and local relevance.

However, several limitations remain. Our evaluation focused on structured MCQs and SAQs, which do not capture the full complexity of clinical encounters. Biomedical models were underrepresented among the top performers, pointing to gaps in current domain adaptation strategies. Finally, while we examined error patterns and prompting effects, further investigation is needed to assess real-world impact, trust calibration, and long-term model behavior in dynamic care settings.

Future work should focus on creating multimodal, context-aware LLMs trained with diverse, regionally grounded datasets. In parallel, regulatory frameworks must evolve to support responsible deployment, ensuring that LLMs complement rather than compromise clinical expertise, particularly in LMICs where the margin for error is narrow and the need for innovation is urgent.

## Methods

### Dataset

We utilized a curated subset of the AfriMed-QA benchmark comprising 3,910 expert-authored vignette-style multiple-choice questions (MCQs) ^16^. These questions were sourced from 20 African medical school professors across five countries—Nigeria, Ghana, Kenya, Malawi, and South Africa—as part of a structured data collection initiative targeting specialty-level clinical knowledge. Each question includes detailed metadata, such as specialty, author role, and clinical domain.

The question set spans 32 medical specialties, with a strong representation from surgical and high-impact specialties relevant to LMIC settings, including General Surgery, Obstetrics & Gynecology, Pediatrics, Emergency Medicine, Neurosurgery, and Infectious Disease. Country-wise, Nigeria, Ghana, and South Africa contributed the largest number of expert MCQs. The complete list of specialties, mapping to Surgical or Medical and count of questions per speciality is provided in Supplementary Material Table 1A and 1B.

### Large Language Models (LLMs)

#### LLM Selection

We evaluated a total of 42 LLMs covering a spectrum of model families and configurations:

- **Size**: ranging from 2 billion to over 500 billion parameters.
- **Access**: including both open-source (e.g., LLaMA-3, Mistral, Phi-3) and proprietary/closed models (e.g., GPT-4o, Claude-3, Gemini Ultra).
- **Domain**: both general-purpose LLMs and domain-specific biomedical models such as JSL-MedLlama, OpenBioLLM, and Meditron.

This diversity allowed us to compare performance across architectures, access models, and optimization strategies in both generalist and specialist domains. The complete list of LLMs is provided in Supplementary Table 2

#### LLM Inference Experiments

All model evaluations were conducted using a reproducible, uniform inference pipeline. We ran open-source models using Hugging Face’s transformers library or Google Model Garden on cloud GPUs (NVIDIA A100, T4, or L4), while closed models were accessed via proprietary APIs with default hyperparameters, standardized token limits, temperature, and generation length to ensure consistency across LLM families.

#### Prompting Approaches

To evaluate the impact of prompt engineering on model performance, we applied four prompting strategies to a representative subset of closed and open three models (GPT-4o, LLaMA3-70B, Gemma-3-27B):

- **Base prompt**: direct question with minimal context.
- **Base prompt + Few-shot:** direct question with 3 in-context examples
- **Instruct prompt**: persona-based roleplay framing the LLM as a skilled African physician.
- **Instruct + Few-shot prompt**: question preceded by 3 in-context examples.

### Evaluation Measures

#### Quantitative Evaluation

- **MCQs**: Model performance was computed using exact-match **accuracy**, defined as the proportion of questions where the LLM-selected answer matched the gold standard.
- **Statistical Analysis**: We used the Student’s T-test to assess statistical significance in model performance across medical and surgical specialties.

#### Qualitative Evaluation

##### Human Evaluation Setup

To capture nuance beyond automated metrics, we conducted clinician-led human evaluations using a web-based double-blind interface. We recruited 58 board-certified physicians and medical faculty from 10 countries, trained on a structured rubric and blinded to whether responses came from a model or human.

### Error Analysis

#### Manual Model Error Review and Error Coding

To analyze the nature of LLM errors, we selected five models—GPT-4o, Claude-3.5, Gemini-1.5-pro (Ultra), Meta LLaMA3-70B, and Llama-405B—representing closed and open large and small LLMs, across a spectrum of performance tiers. Of those five models, For each specialty, a team of 6 clinicians manually reviewed 10 correct and 10 incorrect question-answer pairs to identify error patterns. An expert panel collected examples of recurring errors and categorized them into thematic groups. That process involved the expert reviewers independently coding the observed errors, followed by group discussions to reach consus on final error categories. These included E0=Prompting Errors E1=Factually Inaccurate Errors, E2=Hallucinations, E3=Outdated Information, E4=Failure to follow guideline/Population-Specific context, E5=Invalid Inference Errors, E6=Incorrect Clinical Reasoning Errors, E7=Inappropriate Management Errors.

| Error Code | Error Type | Definition of Error Type | Example Errors Captured |
| --- | --- | --- | --- |
| E1 | **Factually Inaccurate, or Deviates from Academic Scientific Consensus** | *Provably false answer*<br>The response states an objective medical fact, figure, drug dose, epidemiologic detail, or diagnostic criterion that is provably false | Wrong lab ranges |
| E2 | **Hallucination** | Fabrication, nonsensical output, extraneous, irrelevant, or improbable scenarios | Citing non-existent trials<br>Invented statistic |
| E3 | **Outdated information** | Data or details that are no longer current, valid, or relevant due to the passage of time or changes in scientific knowledge and medical science. It can be information that was once accurate but has since become obsolete or superseded by newer, more accurate information. | Obsolete drugs treatments |
| E4 | **Failure to follow Local Guidelines / Population-Specific Context** | The response ignores or contradicts *population-specific* factors (age, pregnancy, comorbidities, local antimicrobial resistance, regional protocols, health-system constraints) or fails to follow the **most relevant guideline** for that setting. | Choosing wrong medication for specific populations (Peds, Pre-Post Partum), ignores weight-based dosing in paediatrics; Suggests unavailable test in low-resource clinic; Chooses wrong answer due to variance in regional epidemiology |
| E5 | **Invalid inference** | An invalid inference is an argument where the conclusion is wrong, the model picked a wrong final answer, even if the rationale or prior logic was true. In simpler terms, the conclusion is false even if the supporting information is correct |  |
| E6 | **Incorrect Clinical Reasoning** | The model shows faulty logical chains, misinterprets the question stem, or provides an explanation inconsistent | Contradicts own rationale statement (“because the virus is bacterial...”); |
|  |  | with the chosen answer. Includes cases where the **answer is correct but the rationale is wrong.** | Mismatches lab units; Misreads negative–positive question stem phrasing |
| **E7** | **Inappropriate Management** | The **next step**, treatment, or disposition contradicts accepted care standards *even if the factual recall is correct*. Emphasis on *actionable harm* (drug, dose, urgency). | Advises wrong order of drug use despite accuracy; Omits emergency referral / management |
| **E0** | **Prompting Error** | *Due to prompt design, model selects only 1 answer, when multiple correct answers exist* |  |

#### Counterfactual Testing

To test model robustness and bias, we conducted a **counterfactual analysis** on 4 high-performing LLMs (GPT-4o, Gemini, Meta AI, DeepSeek) with chat interfaces readily available to clinician evaluators. We perturbed structured variables in vignettes that included patient **age, gender**, or **geographic location** (e.g., “travel to Uganda”) and compared changes in model predictions. This enabled assessment of implicit bias, susceptibility to demographic shifts, and the influence of location-aware training data.

## Data Availability

The AfriMedQA dataset is open-source and is available from huggingface at https://huggingface.co/datasets/intronhealth/afrimedqa_v2

## Code Availability

The code for this analysis is open-source and is made available on GitHub at https://github.com/intron-innovation/AfriMed-QA

