## Supplementary Materials for "Probing the Surgical Competence of LLMs: A global health study leveraging AfriMedQA benchmarks"

Supplementary Material

Table 1A: Specialty mapping to Surgical and Non-surgical domains

| **Specialty** | **Domain** |
| --- | --- |
| **Rheumatology** | Medicine |
| **Nephrology** | Medicine |
| **Endocrinology** | Medicine |
| **Gastroenterology** | Medicine |
| **Other** | Medicine |
| **Pulmonary_Medicine** | Medicine |
| **Pathology** | Pathology |
| **Internal_Medicine** | Medicine |
| **Emergency_Medicine** | Medicine |
| **Psychiatry** | Medicine |
| **Oncology** | Surgery |
| **Neurology** | Medicine |
| **Neurosurgery** | Surgery |
| **Ophthalmology** | Surgery |
| **General_Surgery** | Surgery |
| **Cardiology** | Medicine |
| **Dermatology** | Medicine |
| **Infectious_Disease** | Medicine |
| **Otolaryngology** | Surgery |
| **Urology** | Surgery |
| **Orthopedic_Surgery** | Surgery |
| **Hematology** | Pathology |
| **Pediatrics** | Surgery |
| **Obstetrics_and_Gynecology** | Surgery |

Table 1B: Counts of question per specialty

| **specialty** | **Number of Questions** |
| --- | --- |
| Rheumatology | 85 |
| Nephrology | 79 |
| Endocrinology | 137 |
| Gastroenterology | 130 |
| Other | 32 |
| Pulmonary_Medicine | 103 |
| Neurology | 213 |
| Neurosurgery | 31 |
| General_Surgery | 488 |
| Pathology | 293 |
| Ophthalmology | 104 |
| Cardiology | 172 |
| Internal_Medicine | 71 |
| Urology | 24 |
| Hematology | 100 |
| Psychiatry | 173 |
| Otolaryngology | 58 |
| Dermatology | 29 |
| Emergency_Medicine | 23 |
| Oncology | 38 |
| Orthopedic_Surgery | 51 |
| Infectious_Disease | 184 |
| Obstetrics_and_Gynecology | 660 |
| Pediatrics | 585 |

Table 2: T-test p-values comparing means of MCQ accuracy between medical vs surgical specialties

| **Model** | **P-values** |
| --- | --- |
| Phi-3-medium-128k-instruct | 0.0004 |
| phi4 | 0.0009 |
| gemini-pro-1.5 | 0.0009 |
| Mixtral-8x7B-Instruct-v0.1 | 0.0021 |
| meditron-7B | 0.004 |
| claude-3-haiku | 0.0041 |
| Meta-Llama-3.1-8B-Instruct | 0.0114 |
| claude-3-sonnet | 0.0122 |
| Meta-Llama-3-8B | 0.014 |
| gpt-4 | 0.0155 |
| gemini-2.0-pro-exp | 0.0159 |
| gpt-3.5-turbo | 0.0162 |
| gemma-3-27b-it | 0.0227 |
| gpt-4o | 0.0232 |
| claude-3-5-sonnet | 0.0313 |
| gemma2-2b | 0.0334 |
| Mistral-7B-Instruct-v0.2 | 0.0345 |
| Phi-3-mini-128k-instruct | 0.0459 |
| Mistral-7B-Instruct-v0.3 | 0.0481 |
| JSL-MedLlama-3-8B-v2.0 | 0.0549 |
| claude-3-opus | 0.0622 |
| Llama3-OpenBioLLM-8B | 0.0674 |
| gemini-2.0-flash | 0.0787 |
| claude-3-7-sonnet | 0.0825 |
| qwen-2.5-32b | 0.0828 |
| gpt-4o-mini | 0.0851 |
| gemini-ultra | 0.087 |
| PMC-LLAMA-7B-FT | 0.1001 |
| llama-4-maverick-17b-128e-it | 0.1144 |
| Phi-3-mini-4k-instruct | 0.1501 |
| llama3-405b-instruct-maas | 0.1762 |
| phi-4-mini-4k-instruct | 0.1773 |
| gemma2-27b | 0.1882 |
| gemma2-9b | 0.2512 |
| Llama-3.3-70B | 0.3353 |
| Llama3-OpenBioLLM-70B | 0.4562 |
| OpenAI-o1 | 0.5041 |
| Meta-Llama-3-70B-Instruct | 0.5478 |
| deepseek-distill-qwen32b | 0.715 |

Table 3: Effect of Prompting for selected subset of LLMs

|  | **base** | **base + few-shot** | **Instruct** | **Instruct + few-shot** |
| --- | --- | --- | --- | --- |
| Gpt-4o | 0.8289 | 0.8128 | 0.81432 | 0.821228 |
| gemma-3-27B-instruct | 0.709200 | 0.7084 | 0.7028 | 0.7084 |
| llama-3-70b-it | 0.7749 | 0.7693 | 0.7554 | 0.7693 |
